# Mental health before, during, and after COVID-19 (through 2025) in boys, girls, and gender-diverse youth

**DOI:** 10.64898/2026.09.06.26362161

**Authors:** Olli Kiviruusu, Sebastian Therman, Jukka Lehtonen, Eivind Ystrom, Agnieszka Butwicka, Thorhildur Halldorsdottir, Helga Ask, Jaana Suvisaari

## Abstract

**Background:** Adolescent mental health after the COVID-19 pandemic has been a major public health concern. Studies covering the post-pandemic period remain scarce and rarely include gender-diverse youth. We examined trends in mental health among cisgender, transgender, and nonbinary youth during the pandemic and through 2025.

**Methods:** Participants (N=636,201) were adolescents in grades 8–11 from the Finnish School Health Promotion study between 2019 and 2025, a repeated cross-sectional classroom survey.

Validated questionnaires were used to assess clinical levels of depressive, generalized anxiety and social anxiety symptoms, as well as mental well-being.

**Results:** In 2025 there were 81,145 (48.6%) cisgender girls, 1137 (0.7%) transgender boys, 78,898 (47.2%) cisgender boys, 449 (0.3%) transgender girls, and 4658 (2.8%) nonbinary youth. Clinically significant depressive and generalized anxiety symptoms remained more prevalent in 2025 (reference) compared with pre-pandemic levels across all groups (ORs for 2019, 0.48–0.86; p<0.01). Compared with other groups, symptoms were most prevalent among transgender boys and nonbinary youth. Among cisgender youth, generalized anxiety symptoms increased across the post-pandemic period (ORs for 2021 and 2023, 0.81–0.95; p<0.0001). Nonbinary youth were the only group to show a recovery pattern in depressive, generalized anxiety, and social anxiety symptoms (ORs for 2021, 1.36–1.52; p<0.0001) and demonstrated improved mental well-being after the pandemic (beta for 2021, -0.68; p<0.0001).

**Conclusions:** The results indicate that the elevated levels of mental health problems among adolescents associated with the COVID-19 pandemic have not subsided. They also reveal persistent disparities by gender identity and call for targeted policy action.

## Introduction

Youth mental health has been of increasing concern over the past decade (McGorry et al., 2024). The COVID-19 pandemic further exacerbated negative trends in mental health, as documented in longitudinal studies (Madigan et al., 2023; Bosmans et al., 2025). Findings from the post-pandemic period are mixed: some studies report signs of recovery (Henseke and Schoon, 2025), whereas others indicate no improvement (Kiviruusu et al., 2024), and still others suggest partial recovery depending on the outcome and population subgroup (Haskell et al., 2025). Evidence extending to 2024—and particularly to 2025—remains scarce. One recent Italian survey of 11–19-year-olds found no change in anxiety symptoms and a decline in depressive symptoms between 2021 and 2025 (Barbieri et al., 2026). Although informative, the study lacked pre-pandemic baseline data.

During the COVID-19 pandemic, girls, compared with boys, were disproportionately affected by worsening mental health symptoms (Madigan et al., 2023; Kiviruusu et al., 2024). In addition, transgender and nonbinary youth exhibit higher levels of mental health problems compared with their cisgender peers (Kiviruusu et al., 2024) and may have been particularly vulnerable to the adverse effects of the pandemic (Racine et al., 2025). However, our previous report indicated that post-pandemic trends in mental health (2021–2023) showed signs of recovery among gender-diverse youth, whereas no comparable improvement was observed among adolescent cisgender groups (Kiviruusu et al., 2024). Data on later post-pandemic developments regarding gender-diverse youth are lacking.

We present data on mental health among Finnish youth from the pre-pandemic period through 2025, based on a large nationwide survey comprising 636,201 participants and extending to three years after the cessation of COVID-19-related restrictions in Finland. We examine differences across gender identity groups, including cisgender, transgender, and nonbinary youth, and report both mental health symptoms and mental well-being.

## Methods

### Participants

We used data from the School Health Promotion (SHP) study, a nationwide anonymous classroom survey, conducted biennially by the Finnish Institute for Health and Welfare (THL) using total population sampling (THL, 2025). Responding to the survey is voluntary and based on informed consent. The institutional review board of THL has approved the SHP research plan. Participants were 636,201 lower and upper secondary students (grades 8–11) between 2019–2025, mean age 15.7 (SD 1.1) years. The coverage rate in 2025 was 73% among lower secondary, 70% in general, and 38% in vocational upper secondary schools (THL, 2025).

### Measures

The two-item Patient Health Questionnaire (PHQ-2), using a cutoff of ≥3, and the Generalized Anxiety Disorder Scale (GAD-7), using a cutoff of ≥10, were used to measure depressive and generalized anxiety symptoms, respectively (Kroenke et al., 2003; Spitzer et al., 2006). The three-item Mini Social Phobia Inventory (Mini-SPIN) was used to detect symptoms of social anxiety, with a cutoff of ≥6 (Ranta et al., 2012). Positive mental health was assessed using the Short Warwick–Edinburgh Mental Well-being Scale (SWEMWBS) (Stewart-Brown et al., 2009). All measures were administered biennially from 2019 to 2025, except for Mini-SPIN and SWEMWBS, which were not administered in 2019.

Self-reported official gender (boy/girl) and gender identity (boy/girl/both/neither/it varies) were combined for categories cisgender girl (officially girl, identity girl), transgender boy (officially girl, identity boy), cisgender boy (officially boy, identity boy), transgender girl (officially boy, identity girl), and nonbinary youth (identity both, neither, or it varies).

Background factors included grade (8-11), family’s financial situation (good, moderate, poor), living with both parents (yes/no), and family origin (Finnish vs. immigrant), all self-reported by the students. Information on school level (lower secondary school, general upper secondary school, vocational upper secondary school) was retrieved from the SHP database.

### Statistical analysis

After excluding unreliable responses (1.3%) due to implausible response patterns (THL, 2025, p. 26), 628,058 cases remained for the analyses, which were conducted using SPSS Statistics 31.0. Analyses were performed for the total sample and stratified by gender identity. For each outcome, percentages of those above the cutoff (means for the SWEMWBS) were estimated with 95% confidence intervals. Logistic and linear mixed-effects models were used to analyze changes in outcomes. Study year was included as a categorical predictor, with 2025 as the reference year, and school-level random intercepts were specified to account for the clustering of observations within schools. All mixed-effects models were adjusted for background factors, including school level, grade, family’s financial situation, living with both parents, family origin, and models for the total sample were additionally adjusted for gender identity.

## Results

Frequencies and the extent of missing data for gender identity and background factors are presented in Table 1. The majority of participants were cisgender, with proportion ranging between 94.6% and 96.4% across the study period. The proportion of transgender boys ranged from 0.5% (n = 722) to 0.8% (n = 1193) across study years, whereas transgender girls accounted for 0.2% (n = 276) to 0.3% (n = 449). The proportion of nonbinary youth increased from 2.5% (n = 3808) in 2019 to 3.7% (n = 5789) in 2021, peaked at 3.9% (n = 5805) in 2023, and declined to 2.8% (n = 4658) in 2025 (Table 1).

**Table 1.**
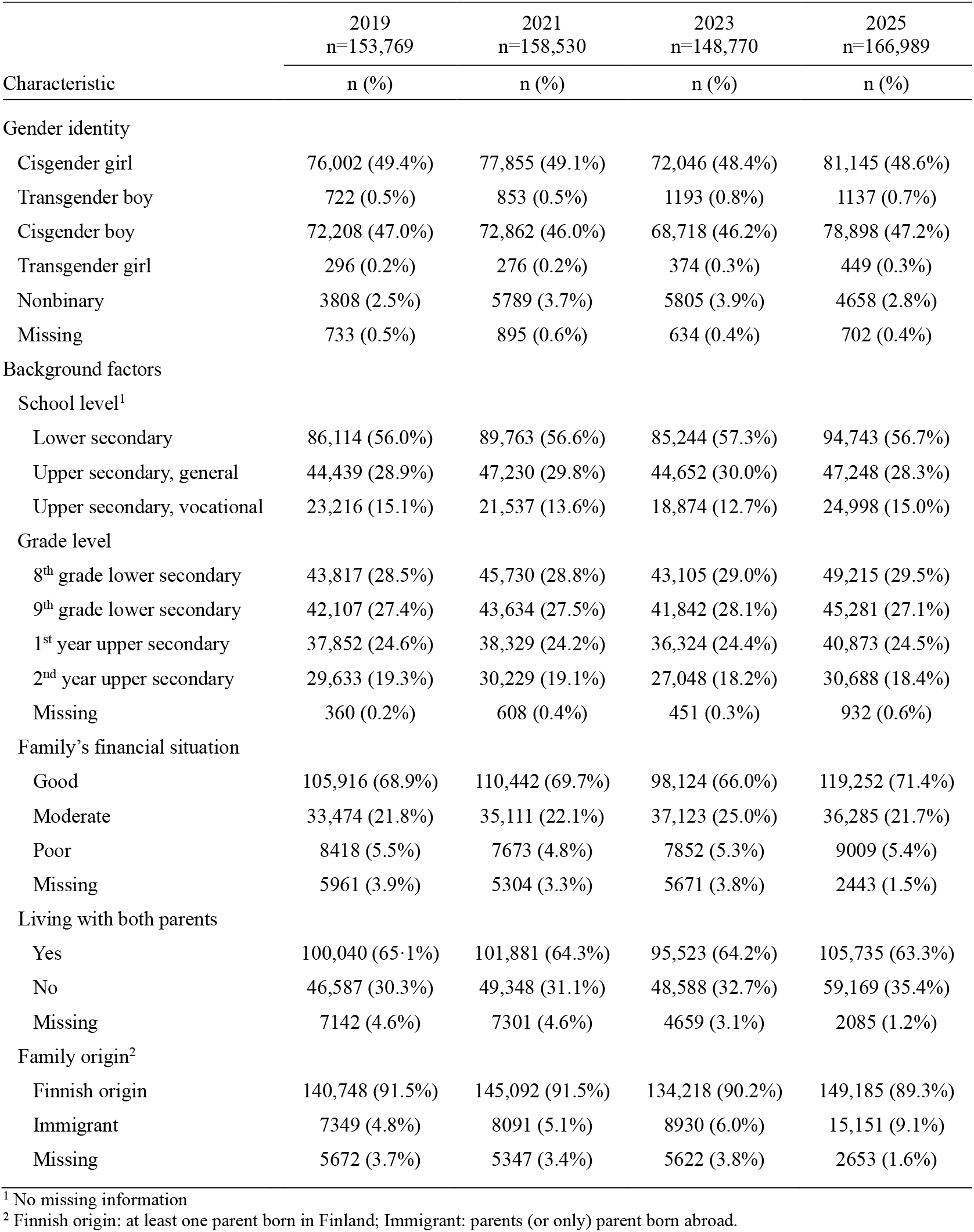
Characteristics of the sample by study year.

|  | 2019<br>n=153,769 | 2021<br>n=158,530 | 2023<br>n=148,770 | 2025<br>n=166,989 |
| --- | --- | --- | --- | --- |
| Characteristic | n (%) | n (%) | n (%) | n (%) |
| Gender identity |  |  |  |  |
| Cisgender girl | 76,002 (49.4%) | 77,855 (49.1%) | 72,046 (48.4%) | 81,145 (48.6%) |
| Transgender boy | 722 (0.5%) | 853 (0.5%) | 1193 (0.8%) | 1137 (0.7%) |
| Cisgender boy | 72,208 (47.0%) | 72,862 (46.0%) | 68,718 (46.2%) | 78,898 (47.2%) |
| Transgender girl | 296 (0.2%) | 276 (0.2%) | 374 (0.3%) | 449 (0.3%) |
| Nonbinary | 3808 (2.5%) | 5789 (3.7%) | 5805 (3.9%) | 4658 (2.8%) |
| Missing | 733 (0.5%) | 895 (0.6%) | 634 (0.4%) | 702 (0.4%) |
| Background factors |  |  |  |  |
| School level<sup>1</sup> |  |  |  |  |
| Lower secondary | 86,114 (56.0%) | 89,763 (56.6%) | 85,244 (57.3%) | 94,743 (56.7%) |
| Upper secondary, general | 44,439 (28.9%) | 47,230 (29.8%) | 44,652 (30.0%) | 47,248 (28.3%) |
| Upper secondary, vocational | 23,216 (15.1%) | 21,537 (13.6%) | 18,874 (12.7%) | 24,998 (15.0%) |
| Grade level |  |  |  |  |
| 8<sup>th</sup> grade lower secondary | 43,817 (28.5%) | 45,730 (28.8%) | 43,105 (29.0%) | 49,215 (29.5%) |
| 9<sup>th</sup> grade lower secondary | 42,107 (27.4%) | 43,634 (27.5%) | 41,842 (28.1%) | 45,281 (27.1%) |
| 1<sup>st</sup> year upper secondary | 37,852 (24.6%) | 38,329 (24.2%) | 36,324 (24.4%) | 40,873 (24.5%) |
| 2<sup>nd</sup> year upper secondary | 29,633 (19.3%) | 30,229 (19.1%) | 27,048 (18.2%) | 30,688 (18.4%) |
| Missing | 360 (0.2%) | 608 (0.4%) | 451 (0.3%) | 932 (0.6%) |
| Family's financial situation |  |  |  |  |
| Good | 105,916 (68.9%) | 110,442 (69.7%) | 98,124 (66.0%) | 119,252 (71.4%) |
| Moderate | 33,474 (21.8%) | 35,111 (22.1%) | 37,123 (25.0%) | 36,285 (21.7%) |
| Poor | 8418 (5.5%) | 7673 (4.8%) | 7852 (5.3%) | 9009 (5.4%) |
| Missing | 5961 (3.9%) | 5304 (3.3%) | 5671 (3.8%) | 2443 (1.5%) |
| Living with both parents |  |  |  |  |
| Yes | 100,040 (65.1%) | 101,881 (64.3%) | 95,523 (64.2%) | 105,735 (63.3%) |
| No | 46,587 (30.3%) | 49,348 (31.1%) | 48,588 (32.7%) | 59,169 (35.4%) |
| Missing | 7142 (4.6%) | 7301 (4.6%) | 4659 (3.1%) | 2085 (1.2%) |
| Family origin<sup>2</sup> |  |  |  |  |
| Finnish origin | 140,748 (91.5%) | 145,092 (91.5%) | 134,218 (90.2%) | 149,185 (89.3%) |
| Immigrant | 7349 (4.8%) | 8091 (5.1%) | 8930 (6.0%) | 15,151 (9.1%) |
| Missing | 5672 (3.7%) | 5347 (3.4%) | 5622 (3.8%) | 2653 (1.6%) |

<sup>1</sup> No missing information<sup>2</sup> Finnish origin: at least one parent born in Finland; Immigrant: parents (or only) parent born abroad.

Depressive symptoms and generalized anxiety increased markedly from pre-pandemic levels to 2021 across all groups (Table 2). In the total sample, the prevalence of clinically significant depressive symptoms rose from 14.8% to 20.7%, and generalized anxiety from 12.6% to 19.3%. Clinically significant symptoms were most prevalent among transgender boys and nonbinary youth, followed by transgender and cisgender girls, while cisgender boys reported the lowest levels. Social anxiety symptoms were the most common, affecting over one-third of the total sample and nearly two-thirds of transgender boys above the established cutoff.

**Table 2.** Prevalence of clinically significant depressive (PHQ-2 ≥ 3), generalized anxiety (GAD-7 ≥ 10), and social anxiety symptoms (Mini-SPIN ≥ 6), and means of mental well-being (SWEMWBS) scores by gender identity and study year.

|  | Cisgender girl |  | Transgender boy |  | Cisgender boy |  | Transgender girl |  | Nonbinary |  | Total |  |
| --- | --- | --- | --- | --- | --- | --- | --- | --- | --- | --- | --- | --- |
|  | % | 95% CI | % | 95% CI | % | 95% CI | % | 95% CI | % | 95% CI | % | 95% CI |
| PHQ-2 $\geq 3$ | | | | | | | | | | | | |
| 2019 | 19.5 | 19.2–19.8 | 39.9 | 36.2–43.5 | 8.1 | 7.9–8.3 | 26.3 | 21.2–31.5 | 40.5 | 38.9–42.1 | 14.8 | 14.6–15.0 |
| 2021 | 27.5 | 27.2–27.9 | 51.8 | 48.4–55.2 | 10.3 | 10.1–10.5 | 36.8 | 31.0–42.6 | 54.5 | 53.2–55.8 | 20.7 | 20.5–20.9 |
| 2023 | 29.4 | 29.1–29.8 | 52.9 | 50.0–55.8 | 10.4 | 10.2–10.6 | 35.5 | 30.5–40.5 | 48.1 | 46.8–49.4 | 21.6 | 21.4–21.8 |
| 2025 | 28.3 | 29.0–28.6 | 49.5 | 46.6–52.5 | 10.5 | 10.3–10.7 | 38.1 | 33.4–42.8 | 44.7 | 43.3–46.2 | 20.6 | 20.4–20.8 |
| GAD-7 $\geq 10$ | | | | | | | | | | | | |
| 2019 | 18.8 | 18.6–19.1 | 32.6 | 29.2–36.1 | 4.6 | 4.5–4.8 | 19.9 | 15.3–24.5 | 32.3 | 30.8–33.8 | 12.6 | 12.4–12.7 |
| 2021 | 28.5 | 28.2–28.8 | 45.3 | 42.0–48.7 | 6.8 | 6.6–6.9 | 29.6 | 24.1–35.1 | 48.6 | 47.3–49.9 | 19.3 | 19.1–19.5 |
| 2023 | 31.2 | 30.8–31.5 | 51.1 | 48.3–54.0 | 7.0 | 6.8–7.2 | 28.0 | 23.3–32.6 | 44.8 | 43.5–46.1 | 20.7 | 20.5–20.9 |
| 2025 | 31.7 | 31.4–32.1 | 48.1 | 45.1–51.0 | 8.3 | 8.1–8.5 | 32.1 | 27.6–36.6 | 41.6 | 40.1–43.1 | 21.2 | 21.0–21.4 |
| Mini-SPIN $\geq 6$<sup>1</sup> | | | | | | | | | | | | |
| 2019 | na |  | na |  | na |  | na |  | na |  | na |  |
| 2021 | 45.4 | 45.0–45.7 | 63.7 | 60.5–67.0 | 20.6 | 20.3–20.9 | 49.6 | 43.6–55.7 | 63.7 | 62.5–65.0 | 34.8 | 34.5–35.0 |
| 2023 | 49.1 | 48.7–49.4 | 65.7 | 62.9–68.4 | 19.6 | 19.3–19.9 | 45.0 | 39.8–50.3 | 61.0 | 59.7–62.3 | 36.1 | 35.8–36.3 |
| 2025 | 47.9 | 47.5–48.2 | 64.8 | 62.0–67.7 | 21.3 | 21.0–21.6 | 45.6 | 40.8–50.5 | 55.2 | 53.7–56.7 | 35.8 | 35.5–36.0 |
| SWEMWBS score, mean<sup>1</sup> |  |  |  |  |  |  |  |  |  |  |  |  |
| 2019 | na |  | na |  | na |  | na |  | na |  | na |  |
| 2021 | 21.2 | 21.1–21.2 | 19.2 | 18.9–19.6 | 23.0 | 23.0–23.0 | 20.6 | 20.0–21.2 | 18.9 | 18.8–19.0 | 21.9 | 21.9–21.9 |
| 2023 | 20.8 | 20.7–20.8 | 19.0 | 18.8–19.2 | 22.6 | 22.6–22.7 | 20.2 | 19.7–20.6 | 19.3 | 19.2–19.4 | 21.6 | 21.5–21.6 |
| 2025 | 21.3 | 21.2–21.3 | 19.4 | 19.2–19.6 | 22.8 | 22.8–22.9 | 20.5 | 20.0–21.0 | 19.5 | 19.4–19.7 | 21.9 | 21.9–21.9 |

Abbreviations: PHQ-2, Patient Health Questionnaire-2; GAD-7, Generalized Anxiety Disorder Scale-7; Mini-SPIN, Mini Social Phobia Inventory; SWEMWBS, Short Warwick–Edinburgh Mental Well-being Scale.
<sup>1</sup> Data not available for 2019

Positive mental health scores were highest among cisgender boys and lowest among nonbinary youth and transgender boys (Table 2). Overall, changes in mean positive mental health scores across the study period were modest.

Adjusted models showed that clinically significant depressive symptoms remained more prevalent in 2025 (reference year) than in the pre-pandemic year 2019 across all gender identity groups (ORs, 0.54–0.86; all statistically significant) (Table 3). Similarly, generalized anxiety levels were higher in 2025 than in 2019. Notably, among cisgender youth, prevalence continued to increase throughout the post-pandemic period from the already heightened levels observed during the pandemic.

**Table 3.**
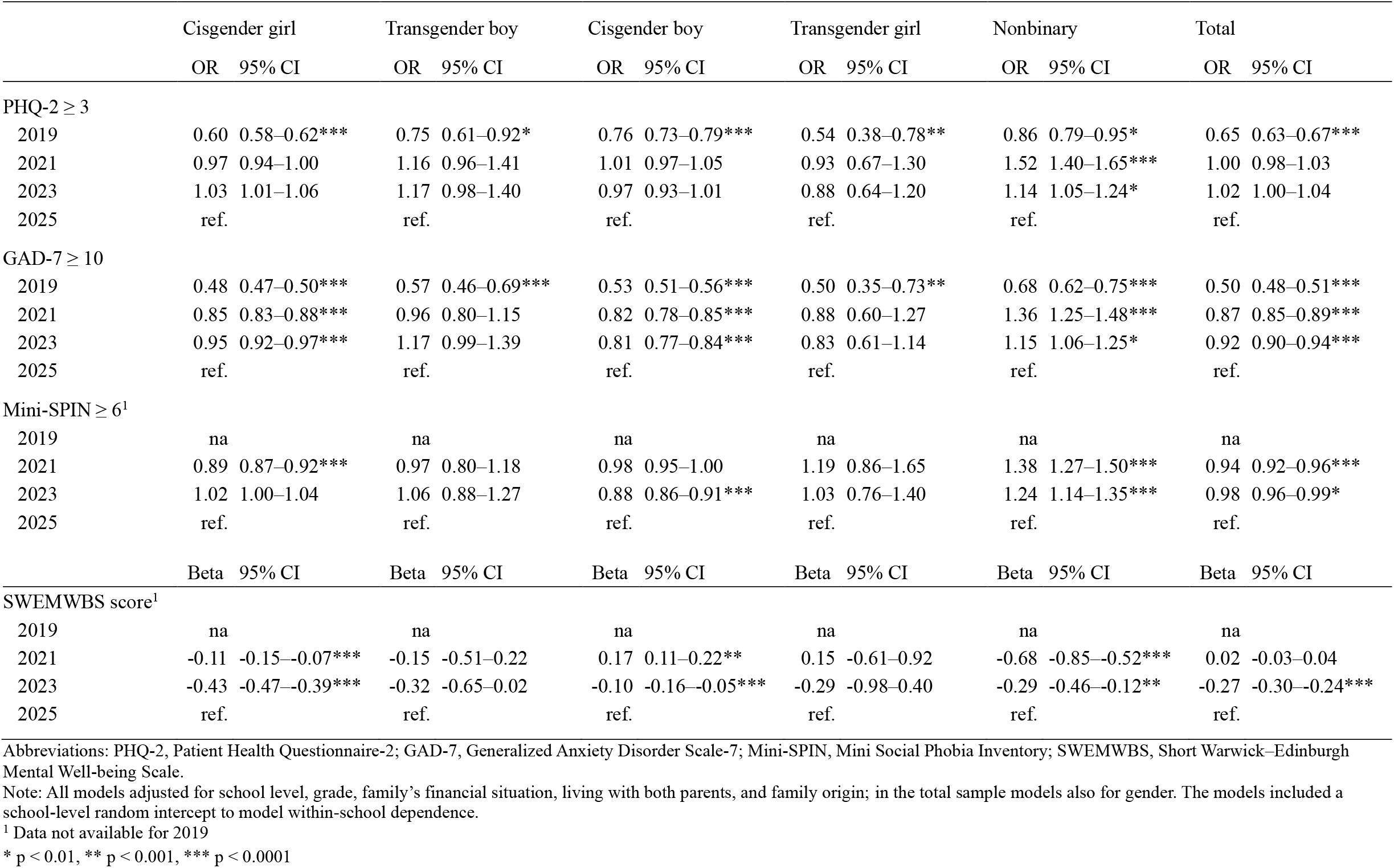
Odds ratios for year from generalized mixed models on clinically significant depressive (PHQ-2 ≥ 3), generalized anxiety (GAD-7 ≥ 10), and social anxiety symptoms (Mini-SPIN ≥ 6), and regression estimates from linear mixed models on mental well-being (SWEMWBS) scores by gender identity.

Nonbinary youth were the only group to show a clear recovery pattern in both depressive symptoms (OR for 2021 = 1.52, 95% CI 1.40–1.65) and generalized anxiety symptoms (OR for 2021 = 1.32, 95% CI 1.25–1.48) after the pandemic peak in 2021 (Table 3). A similar pattern was observed for depressive symptoms among transgender boys, although the results were not statistically significant.

In the total sample, social anxiety increased from 2021 to 2025 (OR for 2021 = 0.94, 95% CI 0.92–0.96) (Table 3). Again, nonbinary youth were the only gender identity group to show a significant improvement, with lower levels of social anxiety in 2025 compared with 2021 (OR for 2021 = 1.38, 95% CI 1.27–1.50).

Positive mental health improved in the total sample from 2023 to 2025 (Table 3). Nonbinary youth and cisgender girls showed statistically significant improvements in mental well-being throughout the post-pandemic period.

## Discussion

We reported post-pandemic mental health trends among Finnish adolescents from 2019 to 2025. One of the most striking findings was that, even three years after the lifting of COVID-19-related restrictions, there was little evidence of recovery in population-level mental health symptoms among Finnish youth. Among cisgender girls and boys, the proportion scoring above the cutoff for generalized anxiety symptoms has continued to increase even after the peak of the pandemic. The persistently high symptom levels are unlikely to be solely attributable to the pandemic; other concurrent crises and broader societal trends have likely contributed (McGorry et al., 2024). Moreover, an increasing trend in mental health symptoms had already been observed before the pandemic, although the additional impact of COVID-19 on top of this trend appeared substantial (Kiviruusu et al., 2023). Regardless of the underlying causes, the high prevalence rates remain a matter of concern. They constitute a significant public health issue and place sustained pressure on mental health services that are already struggling to meet current demand.

Our results indicate that gender-diverse youth showed the most favorable post-pandemic developments. The present analysis extends our earlier study (Kiviruusu et al., 2024) by examining nonbinary adolescents as a distinct group rather than combining them with other gender-diverse youth. The findings suggest that the observed post-pandemic improvement in prevalence rates among gender-diverse youth was largely driven by nonbinary adolescents. Among this group, the positive trend has also continued. Future studies are needed to understand why the positive developments occurred specifically in this group.

Despite these encouraging trends, gender-diverse youth reported by far the highest levels of mental health problems and the lowest levels of positive mental health. This pattern is consistent with previous research and applies to both sexual and gender minority groups (Sares-Jäske et al., 2023; Racine et al., 2025). These populations are disproportionately exposed to discrimination, harassment, and bullying (Sares-Jäske et al., 2023). Such findings highlight persistent inequalities and structural discrimination in society. Addressing these challenges extends beyond the capacity of health services alone and requires broader societal action. One promising intervention target, particularly relevant for adolescents, is the improvement of school climate and safety (Ancheta et al., 2021).

When interpreting our findings, it should be noted that the data are based solely on self-reports and capture symptoms rather than clinical diagnoses. Response rates were reasonably good overall, although considerably lower among vocational upper secondary students. This limitation warrants caution when generalizing the findings to this population. Furthermore, our analyses are based on repeated cross-sectional surveys and therefore do not allow the examination of individual-level changes.

Despite these limitations, we consider the findings compelling. The observed increasing trends are concerning and warrant continuous monitoring. Moreover, the pronounced disparities between gender identity groups call for urgent societal action.

## Data Availability

Finnish Institute for Health and Welfare (THL) has collected the data and has the rights for the data. THL produces public statistical reports and interactive reports, but the data is confidential and thus not publicly available. Researchers can apply the data from Findata (https://findata.fi/en/) or the 2019 and 2025 data from Finnish Social Science Data Archive (https://www.fsd.tuni.fi/en/).

https://www.fsd.tuni.fi/en/

